# Development of a Video-based Evidence Synthesis Knowledge Translation Resource: Applying a User-Centred Approach

**DOI:** 10.1101/2021.03.19.21253944

**Authors:** Cristian Deliv, El Putnam, Declan Devane, Patricia Healy, Amanda Hall, Sarah Rosenbaum, Elaine Toomey

## Abstract

**Background:** People of all ages and walks of life are bombarded with health claims from an array of sources. An understanding of evidence synthesis is important for people to make truly informed healthcare decisions. There is an increasing focus on the use of knowledge translation resources within healthcare; however, the development of these resources has often been poorly described or studied.

**Objectives:** This study employs a user-centred approach to develop a video animation resource to explain the purpose, use and importance of evidence synthesis to the general public regarding healthcare decision-making.

**Methods:** We employed a user-centred approach to developing a spoken animated video that could explain evidence synthesis to a public audience, conducting several cycles of idea generation, prototyping, user-testing, analysis and refinement. Six researchers with expertise in evidence synthesis and knowledge translation resource development gave input on the key messages of the video animation and informed the first draft of the storyboard and script. Seven members of the public provided feedback on this draft through Think-aloud interviews, which we used to develop a video animation prototype. Seven additional members of the public participated in Think-aloud interviews while watching the video prototype. In addition to interviews, participants completed a questionnaire that collected data on perceived usefulness, desirability, clarity and credibility. One experienced patient and public involvement (PPI) advocate also provided feedback on the script and prototype. At the end of each feedback cycle, we assimilated all data and made necessary changes, resulting in a final, rendered version of the animation video.

**Results:** Researchers identified the initial key messages for the SAV as 1) the importance of evidence synthesis, 2) what an evidence synthesis is and 3) how evidence synthesis can impact healthcare decision-making. Using guidance and feedback from members of the public, we produced a three-and-a-half-minute video animation that members of the public rated 9/10 for usefulness, 8/10 for desirability, 8/10 for clarity and 9/10 for credibility. The video was uploaded on YouTube (https://www.youtube.com/watch?v=nZR0xQmZVQg) and has been viewed over 5500 times to date.

**Conclusions:** Employing a user-centred approach, we developed a video animation knowledge translation resource to explain evidence synthesis to the general public that was assessed as useful, desirable and clear by its intended target audience. This study describes the structured and systematic development of this knowledge translation resource and how key stakeholders and end-users informed the final output.

## Introduction

People of all ages and walks of life are bombarded with varying health claims from an array of sources. This is often the information that the public bases its healthcare decisions on [1], yet many of these claims are not reliable and members of the public can find it challenging to assess the reliability of these claims [2, 3]. In addition to this, people’s beliefs about and use of treatments with limited evidence may be harmful. For example, in 2018, a largescale study by Johnson et al. found that patients who received complementary medicine were more likely to refuse conventional cancer treatment and also had a two-fold greater mortality risk compared with no complementary medicine use [4].

Ensuring that healthcare decisions made at all levels (e.g. by policymakers and healthcare managers, practitioners, patients and members of the public) are based on evidence has never been so important. Recent technological advances have led to the availability of vast amounts of health information; however, the sheer quantity of information available can be overwhelming and information overload has been identified as an issue for both clinicians [5] and members of the public alike [3]. Evidence synthesis plays a crucial role in ensuring that healthcare is based on relevant, high quality and up-to-date evidence to optimise health outcomes [6]. Evidence synthesis is an essential way of providing a balanced and comprehensive critical appraisal of all of the available evidence on a topic and is important for stakeholders, including healthcare professionals, policy-makers and members of the public, including patients and their caregivers.

Knowledge translation, or “KT”, is the ‘synthesis, dissemination, exchange and ethically sound application of knowledge to improve health, provide more effective health services and products and strengthen the health care system’ [7]. KT plays a key role in ensuring that research is available and accessible to the public clearly and concisely. Previous research has shown that KT interventions can have beneficial effects on the provision of evidence-based care and patient knowledge and function [8, 9]. Video animations as explanatory resources have been previously shown to effectively communicate complex health information in audiences with different health literacy levels [10]. Spoken animated videos or video animations, in particular, are one of the most impactful ways of conveying information and show high levels of knowledge gain and retention over long periods [11].

KT resources that explain evidence synthesis to a general audience, i.e. not aimed at researchers, are somewhat limited. KT efforts have predominantly focused on increasing awareness and understanding among targeted populations regarding specific clinical topics, disease areas and potential treatments, rather than explaining methodological aspects of healthcare research that are relevant to the general public, such as the importance of evidence synthesis [12-14]. In addition, there is a distinct lack of evidence regarding the development of existing KT resources and how to incorporate an evidence-based approach, and a limited number of examples exist describing this process [15].

Meaningful patient and public involvement (PPI) plays a vital role in ensuring the relevance and potential impact of health research [16, 17]. However, PPI has predominantly been utilised in clinical-based research to date, with less attention to its application in implementation or methodological research [18]. In one recent example from Uganda, a team developed KT resources through PPI approaches. These resources were not only found to be highly effective in a large trial [19], but their success was in part traced back to the user-centred development approach [20].

This study aimed to develop a spoken video animation to explain evidence synthesis and its importance to a public audience by applying a user-centred approach, drawing on cycles of feedback from members of the public to create a useful, usable, understandable, credible and desirable resource.

## Methods

### Ethical approval

As the study aimed to co-produce the KT resource in collaboration with researchers and members of the public, an ethics exemption was granted by the Galway University Hospital Research Ethics Committee. All members of the public provided written or verbal informed consent prior to their involvement.

### Study design

We employed a user-centred approach to determine the format, content and structure of the video animation (Figure 1). User-centred design entails an iterative design cycle with multiple steps: idea generation, prototyping, user-testing and analysis and refinement in collaboration with key stakeholders and end knowledge users, and has been previously employed to develop research resources with input from members of the public [19, 21].

**Figure 1:**
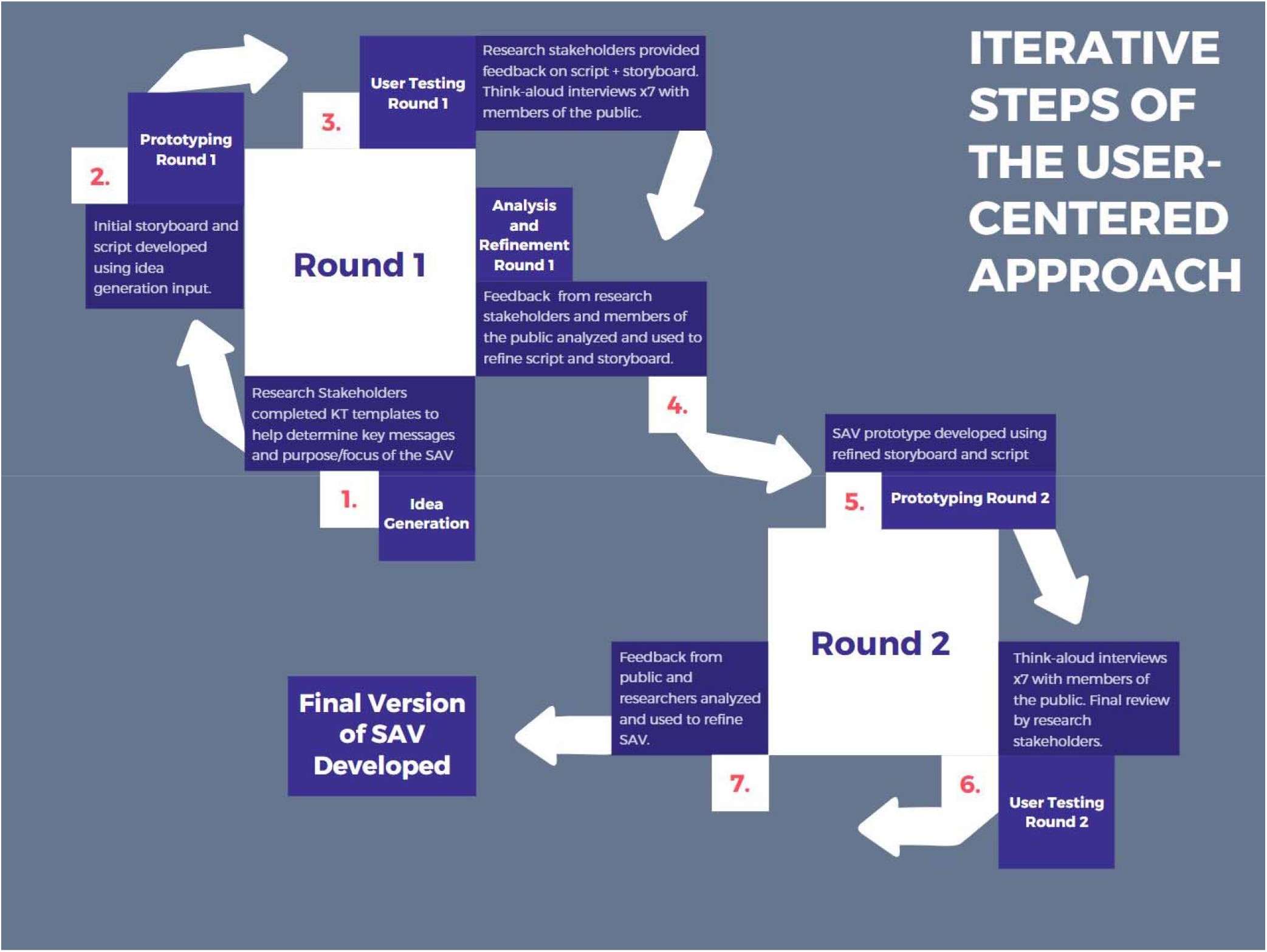
Iterative steps of the user-centred approach SAV: Spoken Animated Video, KT: Knowledge translation

Feedback was obtained during the user-testing through concurrent “Think Aloud” interviews and a questionnaire. Concurrent Think Aloud interviews involve observing people as they engage with a prototype of the product and listening to them speak aloud any words in their mind as they engage, with participants acting as quasi-researchers [22]. This allows for feedback to be honest, clear and immediate.

After the interviews, members of the public completed a questionnaire previously developed by Nsangi et al. [19], which was based on Morville’s honeycomb model of user experience [23]. Specifically, people rated the prototype on a scale of 1-10 for specific facets of the honeycomb model about usefulness, desirability, valuableness, clarity and/or credibility (1 being less useful, 10 being more useful). Interviews and questionnaires were conducted by CD in-person or via telephone, depending on individual preference.

#### Idea generation

The main aim of the Idea Generation stage was to determine the key messages and purpose of the video animation. We used the “Knowledge Translation (KT) Planning Primer” template from the Public Agency of Canada [24] to structure this process. Briefly, the template helps researchers to document their intended audience, objectives, main messages, resource format, development and delivery process, dissemination, available resources, main barriers and desired impact for the project. First, ET and CD completed the template sections referring to the intended audience, objectives, format, development and delivery process and resources. Next, using purposive sampling, authors identified six researchers from Ireland, the UK, Norway and Australia who had combined research and practice expertise in evidence synthesis and knowledge translation. Researchers received the partially completed KT template to review before participating in an unstructured one-to-one telephone discussion with CD to provide feedback. We particularly sought input regarding the main messages of the animation, ideas for dissemination, potential barriers and desired impact, and any general thoughts on the project or other components of the KT template. CD recorded researchers’ feedback directly into the KT template during the telephone discussions. Subsequently, it collated input from all researchers and entered it into the relevant section of the KT template. ET reviewed this synthesised input before moving to the next stage of prototype development. Any uncertainties or conflicting opinions between researchers were discussed between CD, ET and DD.

#### Prototyping – Round One

Using the researchers’ feedback in the Idea Generation stage, we created a first version of the narration script and rough storyboard outline. The storyboard consisted of sketches of each scene (7 scenes in total) and verbal descriptions of approximate visuals scene-by-scene.

#### User Testing – Round One

We recruited members of the public through convenience sampling and snowball/word of mouth to achieve a variety of age and gender. All had limited previous experience of evidence synthesis or health research. People were not offered any incentives for their involvement. User testing sought to obtain feedback on the prototype and determine necessary changes to the narration script, the storyboard, and the animation’s visual and aesthetic style. People were also shown four types of animation styles - felt stop motion, 2D vector, paper cut-out look and whiteboard drawing. During the Think Aloud interviews, members of the public were asked for their first impressions of the script, the visuals accompanying the script, and their impressions of the ‘main messages’ of the video. All were given identical instructions to “think their thoughts aloud and say anything on their mind at any point while interacting with the resource”. Voiced thoughts and comments were documented by CD as facilitator notes (CD).

After the interviews, people were asked to rate the script and storyboard on a scale of 1-10 for specific facets of the honeycomb model about usefulness, desirability, valuableness and credibility. They were also asked to identify any potential missing aspects, content that should be removed, or general suggestions for improving the script and storyboard. The complete questionnaire is provided in Appendix 1.

We obtained feedback on the initial script and storyboard from the six researchers and feedback from one specialist PPI advocate (DS) with substantial experience in knowledge translation for the public. We sent a copy of the script and storyboard in Word format to each person for review. All commented on the document using track changes and returned feedback to CD.

#### Analysis and Refinement – Round One

We synthesised and analysed the information and feedback gathered from the user testing with members of the public, researchers and our experienced PPI advocate. Specifically, we extracted all comments suggesting potential edits or changes to the prototype script and visuals from the Think Aloud facilitator notes and questionnaires. We grouped these into similar categories/concepts to identify a list of main suggestions. These suggestions were subsequently discussed for feasibility of changes by ET and CD, and where suggestions differed across contributors or consensus was not reached, a third member of the study team (DD) was consulted.

#### Prototyping – Round Two

Based on the feedback from the first round of user testing, CD created a second version of the script and recorded an accompanying audio narration. With input from EP, CD developed an initial video prototype from the storyboard sketches.

#### User Testing – Round Two

We used snowball sampling to recruit an additional seven members of the public of varying ages and gender for the second round of user testing. As with the first round, CD conducted Think Aloud interviews with all seven members. This time, people were asked to watch the initial video prototype and talk through their first impressions of the video narration, animation and visuals and main messages, with feedback documented by CD as facilitator notes. Stakeholders then completed another questionnaire (Appendix 2) which asked users to rate the video on a scale of 1-10 according to aspects of usefulness, desirability, clarity and credibility with questions slightly modified to be specific to the video animation

This prototype was also sent to the research and our PPI advocate for feedback before the final stage of refinement.

#### Analysis and Refinement – Round Two

We synthesised and analysed information, feedback and answers gathered from the second cycle of user testing and used this to inform the final version of the video animation, including final changes to visuals, script and the speed of narration and animation. In this final stage, we also added further audio aspects to the video, such as the backing music track and sound effects for the animations’ movement to represent the animation’s final look and feel of the animation accurately. Multiple iterations of the video were rendered to finalise aesthetic details such as frame-rate or colour palette. Finally, we equalized the audio levels and completed any final adjustments before rendering the final entire video.

## Results

### Idea Generation

CD and ET completed the sections of the KT planning primer-template as outlined in Table 1. In addition to this, the research stakeholders overall agreed on which main messages should be prioritized: 1) Explaining the importance of evidence synthesis for not making healthcare decisions based on a single piece of research due to potential for skewed or inaccurate views from one study alone, 2) Explaining what evidence synthesis is as the process of combining separate individual studies on a singular topic in a structured and trustworthy way, and 3) Explaining the usefulness of evidence synthesis for the audience, with an example of how it can inform everyday healthcare decision-making. The researchers also identified the importance of including funding information in the video animation and using Cochrane resources to disseminate it.

**Table 1:** KT Planning Primer Template.

| KT Template Section | Completed Sections |
| --- | --- |
| WHO do we want to reach? | Members of the general public with little to no research or professional healthcare background |
| WHY – What are our objectives? | To create a video-based animation resource for our audience, to increase awareness and understanding of what evidence synthesis is (beyond systematic reviews alone) and its importance and role in informing health decision-making amongst the general public |
| HOW – What format will we use? | We will use a video-based Spoken Animation resource. Animation length will be ~3 minutes+/-1 min |
| HOW – How will we develop the resource? | The resource will be co-produced with groups of stakeholders, using a user-centred approach. The first group will comprise researchers with experience of evidence synthesis and knowledge translation to determine the key messages. The second group will comprise members of the public to user test the video development in Think-aloud interviews. An adapted version of Morville's Honeycomb Framework for collecting and analysing data will structure the user-testing. |
| HOW – Choose the delivery | Youtube, social media e.g. Twitter |
| HOW – Assess your resources | One summer student (CD) and two supervisors – evidence synthesis (ET) and technology (EP) |

#### Prototyping - Round One

The initial prototype storyboard consisted of sketches of 7 scenes in total, with a draft narration script and verbal descriptions of approximate accompanying visuals for each scene.

#### User Testing - Round One

Seven members of the public were involved in the first round of user testing (details in Table 2). People rated the storyboard prototype as median 9 (range 8-10) for usefulness, 7 (range 5-10) for desirability, 9 for value (range 6-10) and 9 for credibility (range 8-10). Overall, they commented on the script and visuals favourably. Predominantly, suggestions for improvement pertained to making specific sentences more concise and using simpler language (scenes 2, 5) and having less imagery and activity in the visuals (scenes 2, 4). One person mentioned the need to clarify further the impact of evidence syntheses on everyday life (scene 6). Four people preferred the felt animation style, with one preferring the 2D vector style, and another preferring the whiteboard style.

**Table 2:**
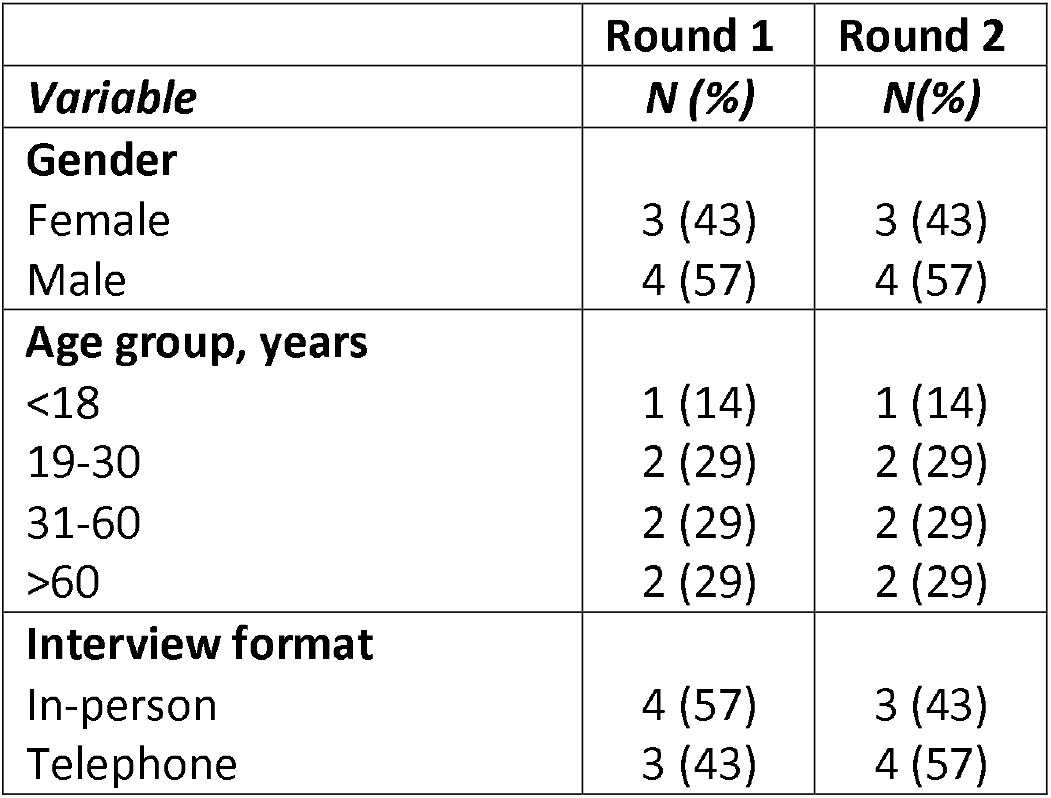
Public Stakeholder Demographics Round 1 and Round 2.

|  | Round 1 | Round 2 |
| --- | --- | --- |
| **Variable** | **N (%)** | **N(%)** |
| **Gender** |  |  |
| Female | 3 (43) | 3 (43) |
| Male | 4 (57) | 4 (57) |
| **Age group, years** |  |  |
| <18 | 1 (14) | 1 (14) |
| 19-30 | 2 (29) | 2 (29) |
| 31-60 | 2 (29) | 2 (29) |
| >60 | 2 (29) | 2 (29) |
| **Interview format** |  |  |
| In-person | 4 (57) | 3 (43) |
| Telephone | 3 (43) | 4 (57) |

Similarly, the research and PPI stakeholders commented favourably overall on the script and visuals. They provided suggestions in relation to wording (e.g. ‘as opposed to a single research study, say “a single piece of evidence”‘) and ensuring the visuals of the video were inclusive and reflective of diversity (e.g. ‘*a number of people emerge - including women, different races and disabilities, a sense of people working together…Each bring different papers and begin to compare and contrast*’).

#### Analysis and Refinement - Round One

After discussion with ET and DD, CD assimilated the synthesised comments and critiques from the first round of user testing and used them to edit the script, audio narrative and storyboard in order to transition to Round 2 of prototype creation.

#### Prototyping - Round Two

A basic video animation was created from the storyboard sketches. The video was kept simple with basic vector animations (simple linear movement) to allow easy changes further in the production process (Figure 2). The audio narration was added to the animation, and in-sync sound effects were added to the animation to develop a second prototype of the video animation resource for user testing in Round 2.

**Figure 2:**
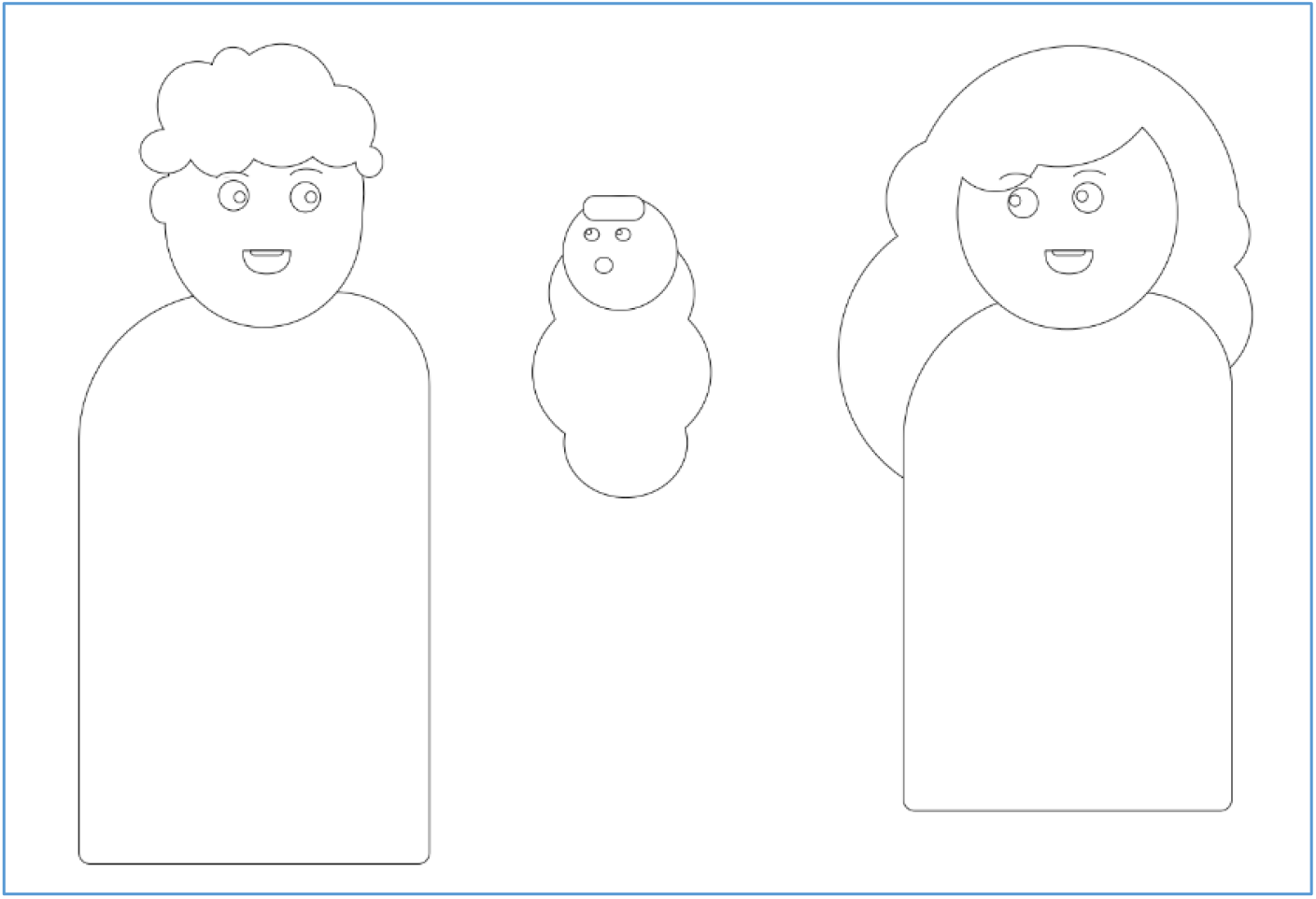
Animation style for Round 2 SAV prototype.

**Figure 3:**
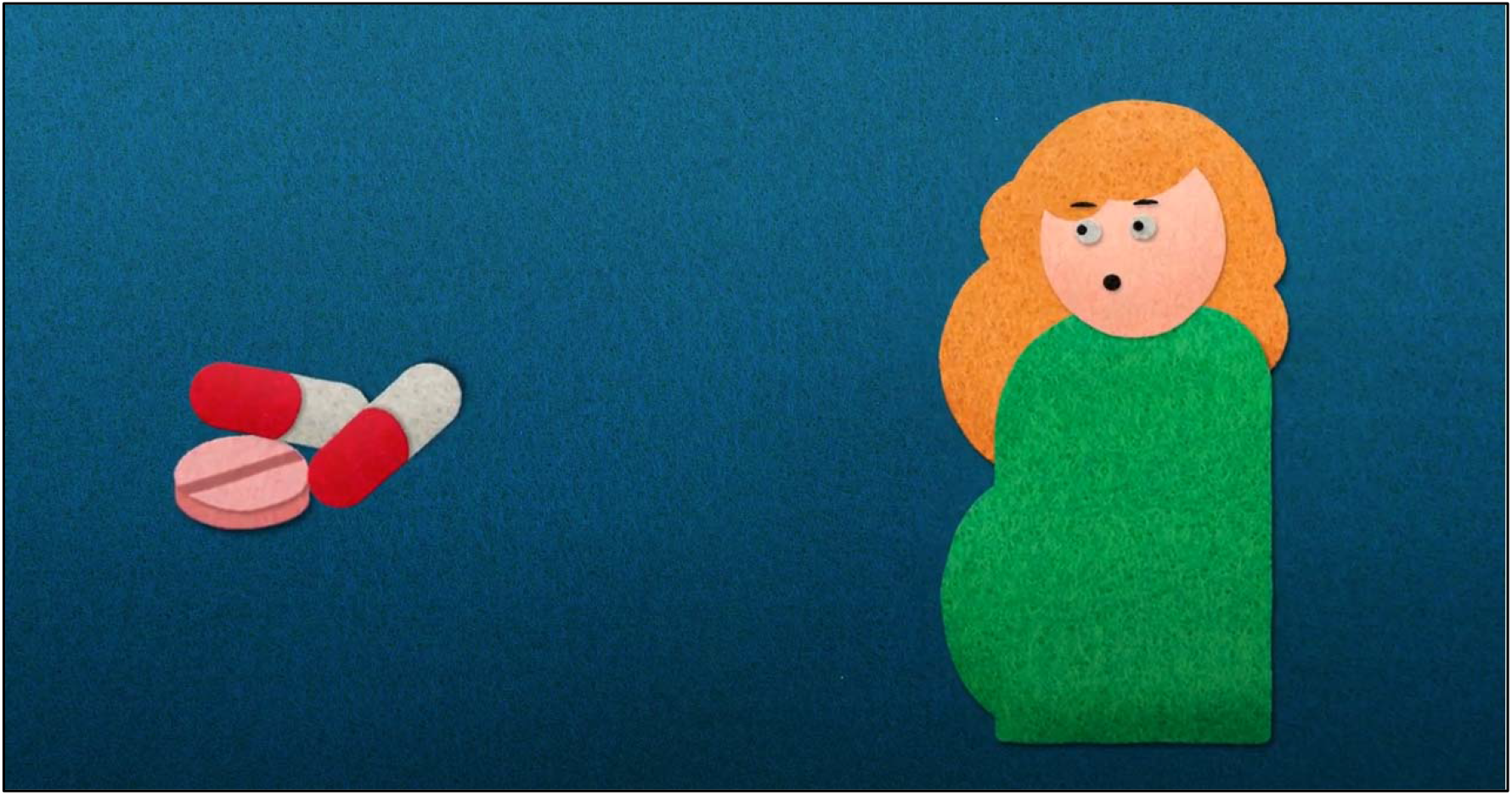
Animation style for final video animation.

#### User Testing - Round Two

Seven additional members of the public were involved in the second round of user testing (details in Table 2). They rated the prototype video animation as median 9 (range 7-9) for usefulness, 8 (range 5-9) for desirability, 8 (range 4-10) for clarity, and 9 (9-10) for credibility. Overall, people liked the visuals and script. Suggestions for editing mostly related to slowing the speed of the visuals and audio in scene 1.

Similarly, researchers and members of the public liked the narration, visuals and script. Minor changes were suggested, such as removing a mnemonic that CD and ET had initially developed to summarise steps for checking the quality of a systematic review to avoid confusion with existing systematic review quality checklists and rephrasing part of the final scene to be more supportive rather than dictatorial (e.g. ‘*hopefully you should feel equipped in using research to make more informed decisions about healthcare’ rather than ‘evidence synthesis can and should directly impact how you make decisions about health care*’).

#### Analysis and Refinement - Round Two

The final version of the video animation was developed using the feedback from the second round of user testing. CD and EP created accurate sound effects, a backing audio track, and intro and outro scenes. CD applied a felt stop-motion animation style in response to user feedback from the first round of user testing. After numerous pre-renderings CD rendered and uploaded the final SAV resource onto the ESI (Evidence Synthesis Ireland) and Cochrane Youtube channels https://www.youtube.com/watch?v=xGdZQNHC8-k&t=1s and https://www.youtube.com/watch?v=nZR0xQmZVQg. At the time of writing, the video has over 1000 views on the ESI Youtube channel and over 4,500 views on the Cochrane Youtube channel. Resource-use was minimal, as this was conducted over 8 weeks as an undergraduate summer student project supported by Evidence Synthesis Ireland for €2000.

## Discussion

Advances in online technology and social media have seen substantial increases in the amount of health information available to and accessed by the public. While there are obvious benefits to using social media and online resources for mass communication of health information, such tools are typically unregulated and the information shared can be of varying quality and consistency [2]. The recent COVID-19 pandemic has shone a spotlight on the twin issues of misinformation and disinformation, i.e. the inadvertent or deliberate and coordinated spread of misleading and false information, respectively [25]. The severity of this situation has led to the coining of the new term ‘infodemic’ by the World Health Organization as the ‘overabundance of information and the rapid spread of misleading or fabricated news, images, and videos’ [26]. Enabling public members to think critically about health claims is a crucial component in the fight to combat the effects of misinformation and disinformation. Moreover, understanding the value and relative importance of a single study on a topic compared to the synthesised body of evidence for a topic is a key part of this [27]. Developing an online KT resource that explains evidence synthesis to a general public can leverage the advantages afforded by social media and be used to spread accurate and valid health information and help people make more informed decisions.

It has been previously estimated that approximately 85% of biomedical research is wasted [28]. Therefore, to minimise research waste and optimise the investment in KT resources and dissemination strategies, it is important to ensure that these are well developed and evidence-based. However, previous research has shown that such resources are often poorly explored and evaluated [29, 30]. Recent work by Cross et al. involved participation between families and researchers in the area of childhood disability to develop, implement and evaluate a web-based KT resource to promote adoption of the ‘F-words’ concepts [15]. The project used the Knowledge-To-Action Framework [31] to guide this process and culminated in developing a theory-informed resource that was deemed relevant and meaningful to its target audiences. The authors highlighted the positive impact of co-production of the resource but also recommended the use of a structured approach for obtaining feedback for future KT resource development projects. In our project, we applied a user-centred design approach to obtain feedback which had been heavily guided by the methodology used by Semakula et al. [21]. This approach enabled us to collect feedback in a structured and iterative manner, building the resource gradually with the input stakeholders embedded throughout the process. Future work will aim to evaluate further the effectiveness and impact of the video resource using rigorous and robust methodologies.

### Strengths and limitations

A strength of our approach is the involvement of multiple individuals, including researchers with expertise in knowledge translation and evidence synthesis, a patient and public advocate with substantial experience in knowledge translation for the public, and members of the public with less experience in health research and knowledge translation. Although efforts were made to recruit members of the public of varying ages and genders, a limitation of our approach is the use of convenience sampling. This meant that some of the public stakeholders were known to the lead authors (CD and ET), and therefore may have felt some degree of pressure not to be too critical of the resource for fear of offending. Attempts were made to mitigate this by providing clear instructions at the outset before user testing about the importance of honest and critical feedback, with the overall purpose of creating the most user-friendly resource possible.

In addition, owing to the user-centred design approach’s iterative nature, additional user-testing cycles could have been applied with additional users who may have yielded additional insights. However, the costs (in time and resources) of conducting additional feedback cycles need to be weighed against that effort’s likelihood resulting in significant improvements. We deemed that two rounds of testing were sufficient as no new insights were identified during the second round of testing.

## Conclusion

In an age of increasing exposure to health information, misinformation and disinformation, a good understanding of evidence synthesis is crucial for enabling people to think critically about treatment claims and choices. We employed a user-centered approach in conjunction with public, research and PPI stakeholders to produce a knowledge translation resource to explain evidence synthesis to members of the public. The input of stakeholders from multiple backgrounds and iterative revisions based on their feedback was critical for ensuring the development of an appropriate and user-friendly resource.

## Supporting information

Appendix 1 Round 1 interviews_questionnaire

Appendix 2 Round 2 interviews_questionnaire

## Data Availability

The data that support the findings of this study are available from the corresponding author, [ET], upon reasonable request.

## Acknowledgements

The authors would like to thank sincerely our public, PPI and researcher knowledge users who contributed to our resource development. They are (alphabetical order): Allen Conor, Booth Andrew, Carey Ryan, Chapman Sarah, Cunningham Niamh, Deliv Daniel, Deliv Eric, Fahey Sarah, Hall Maura, McCool Grainne, McEntee Joe, Mosenet Ion, Nazarewicz Wiktoria, O’Brien John, O’Brien Mary, Ryan Rebecca, Ryan-Vig Selena, Stewart Derek, Zerkaljis Deniss. Cristian Deliv was supported to work on this project by a Summer Studentship from Evidence Synthesis Ireland.

## CRediT Author Statement

Cris Deliv: Investigation, Resources, Data Curation, Visualisation, Writing - Original Draft, Writing - Review & Editing El Putnam: Software, Methodology, Resources, Writing - Review & Editing. Declan Devane: Conceptualization, Methodology, Resources, Writing - Review & Editing, Funding acquisition. Sarah Rosenbaum: Conceptualization, Methodology, Writing - Review & Editing. Amanda Hall: Methodology, Writing - Review & Editing. Patricia Healy: Conceptualization, Resources, Writing - Review & Editing, Funding acquisition. Elaine Toomey: Conceptualization, Methodology, Investigation, Resources, Writing - Original Draft, Writing - Review & Editing, Supervision, Project administration, Funding acquisition.

## Notes

### Competing Interest Statement

The authors have declared no competing interest.

### Funding Statement

Cristian Deliv was supported to work on this project by a Summer Studentship from Evidence Synthesis Ireland. No other external funding was received for this project.

### Author Declarations

An ethics exemption was granted by the Galway University Hospital Research Ethics Committee

