## Appendix 1 Round 1 interviews_questionnaire for "Development of a Video-based Evidence Synthesis Knowledge Translation Resource: Applying a User-Centred Approach"

**User testing Part 2 – Background Questionnaire**

**Instructions**

- **Fill out the short questionnaire, let us know if you have any problems.**
- **Try to answer the questions honestly.**
- **Summarise your experience, adding any additional comments you may have.**
- **Enjoy the process and thank you very much for participating!**

### Background Questionnaire

**User testing Part 2**

**Confidentiality:** We will keep your feedback confidential. Only the people carrying out this study will know the results and feedback you provide us with. If we cite your feedback in any presentation of this study, you will not be identifiable (although your contribution will be acknowledged more generally).

Please fill in the right-hand column of the questionnaire.

| **Name:** |  |
| --- | --- |
| **Age:** |  |
| **Gender:** |  |
| **How would you categorize your competencies?** | **Systematic review/evidence synthesis expertise**  ☐ Experienced (e.g. researcher or review author)  ☐ Knowledgeable but not highly experienced (e.g. communications staff with some familiarity of systematic reviews)  ☐ No experience or training in systematic reviews or evidence syntheses. |

Please complete this section before moving on.

**Think-aloud Interpretation**

**User testing Part 3.1**

**Instructions**

- **Please interpret the material presented to you and try to “think-aloud”, (voice any of your concerns, thoughts or opinions at any point throughout the interpretation) at any time you see fit. Take your time and try to not overthink, say whatever comes to mind while reading the script and interpreting the visuals.**
- **Try to answer the questions as honestly as possibly, even negative critique is very helpful.**
- **While reading the script and the visuals, if possible read them out loud.**
- **When reading out loud, stop to voice your thoughts after each sentence, or each scene or every few words, whichever is most comfortable for you.**
- **When reading the visual depictions of every scene, try to imagine the scenes in your mind, be as vivid as possible about what you are picturing.**
- **Enjoy yourself, remember that this resource is designed to educate the public in an enjoyable, pleasant manner 😊**

Notes:

Please complete this section before moving on.

**User testing Part 3.2**

**Written Comments**

| **What were your first impressions of the script?** |  |
| --- | --- |
| **What were your first impressions of the visuals accompanying the script?** |  |
| **What would you describe as being the “main messages” of this video?** |  |
| **Please circle or highlight the appropriate number rating below.** | |
| **What is its “usefulness” from 1-10? Does this video fill an existing gap in your opinion? Do people need to be educated on the topics covered in this video?** | 1 2 3 4 5 6 7 8 9 10 |
| **What is its “desirability” from 1-10? How desirable would you deem this video to be? How likely are people to want to watch this? How visually attractive and aesthetically pleasing do the visuals seem?** | 1 2 3 4 5 6 7 8 9 10 |
| **How “valuable” would you deem this video and the information provided in it to be to you? How likely are you to remember this video in the future? Are going to apply the information learned in this video to your healthcare decision-making?** | 1 2 3 4 5 6 7 8 9 10 |
| **How “credible” or trustworthy would you rate the information given in this video to be? From the organisations we’re part of and associated with, and from how the information is presented to you.** | 1 2 3 4 5 6 7 8 9 10 |
| **Summarise your general overall experience reading through the script and storyboard** |  |
| **What did you like best?** |  |
| **What caused you the most problem?** Please indicate what you felt were the main obstacles in understanding the script materials. |  |
| **Were there important items you felt were missing?** |  |
| **Was there content you felt should be excluded?** |  |
| **Do you have other suggestions about how we could change the script and storyboard so they would be more useful for you?** |  |
| **Any other comments?** |  |

**Many thanks for participating!**

**Cristian Deliv**, (Undergrad MD at National University of Ireland Galway)

**Dr Elaine Toomey** (PhD, project supervisor, Associate Director of Cochrane Ireland)

**Dr EL Putnam** (PhD, Lecturer and Programme Director in Huston school of Film and Digital Media)

**Prof Declan Devane** (Director of Cochrane Ireland and Evidence Synthesis Ireland).
